# Quantifying Face Mask Comfort

**DOI:** 10.1101/2021.03.31.21254723

**Authors:** Esther Koh, Mythri Ambatipudi, DaLoria L. Boone, Julia B. W. Luehr, Alena Blaise, Jose Gonzalez, Nishant Sule, David J. Mooney, Emily M. He

**Author notes:** These authors contributed equally (Esther Koh, Mythri Ambatipudi). These authors contributed equally (David J. Mooney, Emily M. He).

## Abstract

Face mask usage is one of the most effective ways to limit SARS-CoV-2 transmission, but a mask is only useful if user compliance is high. Through anonymous surveys, we show that mask discomfort is the primary source of noncompliance in mask wearing. Further, through these surveys, we identify three critical parameters that dictate mask comfort: air resistance, water vapor permeability, and face temperature change. To validate these parameters in a physiological context, we performed experiments to measure the respiratory rate and change in face temperature while wearing different types of commonly used masks. Finally, using values of these parameters from experiments and the literature, and surveys asking users to rate the comfort of various masks, three machine learning algorithms were trained and tested to generate overall comfort scores for those masks. Although all three models tested performed with an accuracy of approximately 70%, the multiple linear regression model also provides a simple analytical expression to predict the comfort scores for any face mask provided the input parameters. As face mask usage is crucial during the COVID-19 pandemic, the ability of this quantitative framework to predict mask comfort is likely to improve user experience and prevent discomfort-induced noncompliance.

## Introduction

To limit the airborne spread of severe acute respiratory syndrome coronavirus 2 (SARS-CoV-2), the World Health Organization advises face mask usage in healthcare and community settings (WHO, 2020). Face masks must have high filtration efficiency, secure fit, and be comfortable over long periods of use in order to help mitigate disease transmission. If masks do not meet acceptable standards for comfort, compliance with donning masks may be low, possibly leading to an increase in viral transmission (Park & Jayaraman, 2020). Surveys of healthcare workers indicate how mask discomfort may eventually decrease the compliance (Baig et al., 2010; Krah et al., 2016; Lee et al., 2020). Few studies, however, have correlated comfort with compliance outside of healthcare settings, where compliance is likely a substantially bigger problem.

Comfort can be defined as the breathability and thermal regulation of a mask (Lee et al., 2020). Breathability can be largely gauged by air resistance while thermal regulation is best described by thermal conductivity (Lee et al., 2020). A mask must be breathable to alleviate the feeling of suffocation for the wearer (Park & Jayaraman, 2020) and thermally conductive to guide heat away from the face and prevent microclimates with extreme heat within masks. As accumulation of moisture within a mask is a common complaint, a comfortable mask should also be highly permeable to water vapor to (1) prevent condensation and (2) wick water to the outer surface where it can evaporate (Lee et al., 2020). While previous studies examined a diverse range of commonly used masks (Fischer et al., 2020), further research is needed to validate comfort level predictions based on measurements of the identified parameters.

Methods of assessing face mask comfort have been developed for settings beyond the medical industry. For instance, to evaluate the ergonomic comfort of respirators among healthcare personnel, a questionnaire with a unique set of 42 components was created (Jazani et al., 2018). Other methods rely on rating scales, often Likert scales that range from 1 to 5 (Foereland et al., 2019). While these methods roughly assess mask comfort, they frequently rely on qualitative judgments. More quantitative methods, such as those to evaluate comfort based on breathability (Choi et al., 2020), only focus on one aspect and do not account for other important factors, e.g., thermal regulation (Lee et al., 2020). A standardized metric that uses multiple parameters to quantitatively evaluate face mask comfort has yet to be defined.

In this paper, we present a broad study of key factors affecting mask comfort and a machine learning algorithm to quantitatively assess comfort. We identified air resistance, water vapor permeability, and face temperature change as key factors that determine overall comfort using surveys of students, faculty, and staff on campus at Harvard University. Tests measuring the respiratory rates and face temperature change of volunteers were performed to validate these factors. Finally, using survey results and known values for our parameters, we developed and trained three machine learning algorithms to predict the comfort of masks with an accuracy of ∼70%.

## Results

### Relationship Between Mask Comfort & Compliance

To examine the relationship between comfort and compliance in mask use, an anonymous survey (n = 205) was conducted on students, staff, and faculty residing on Harvard’s campus during the Fall 2020 semester. Out of 170 respondents, 95.3% reported wearing a mask while walking or commuting around campus (Figure 1A). Out of 171 respondents, 90.1% also wore a mask “always” or “most of the time” in gatherings of 2+ people (Figure 1B).

**Figure 1.**
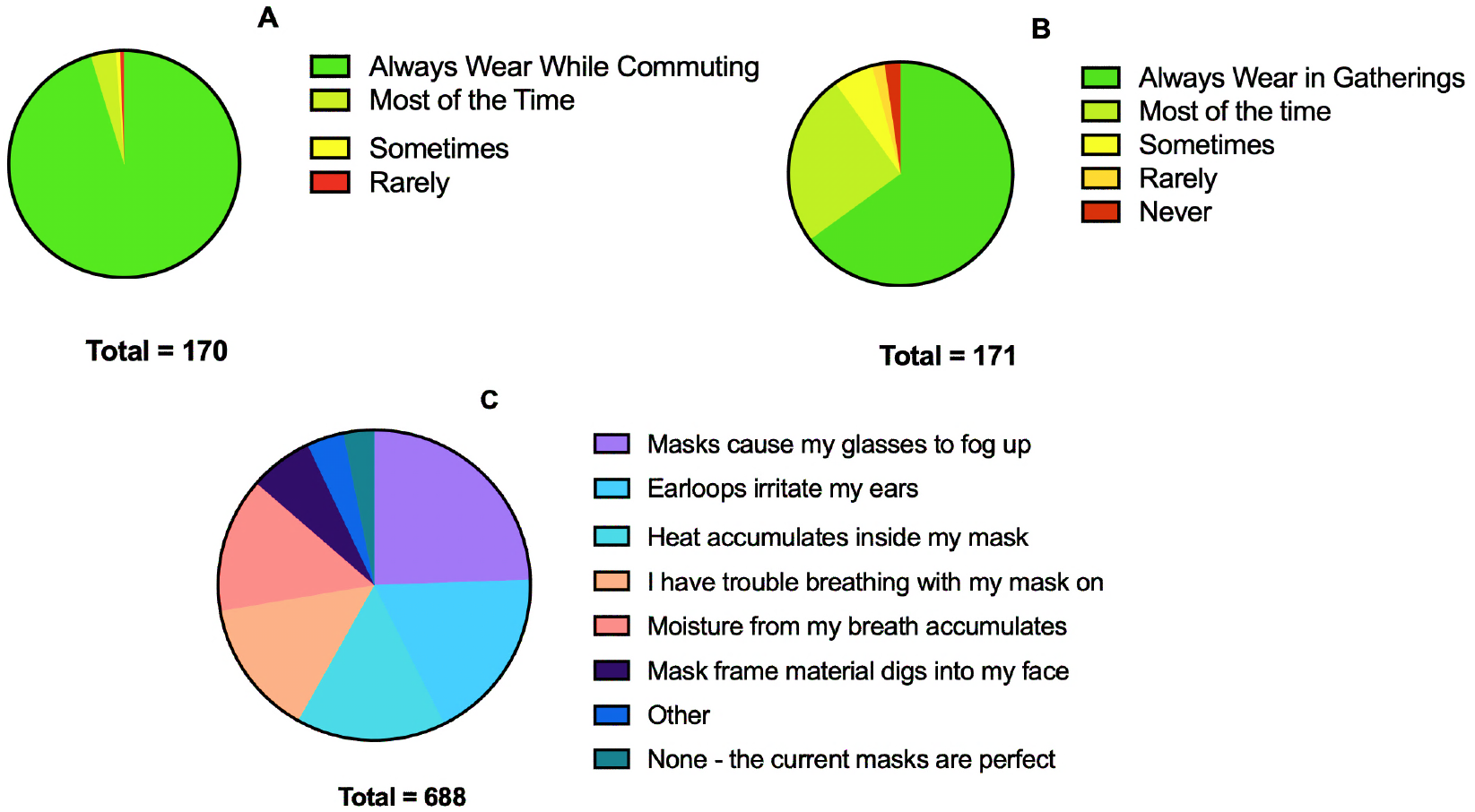
Results of Campus Survey on Mask Use and Comfort: **A)** A large majority of survey participants (95.3%) reported strict adherence to mask wear while walking/commuting on campus. **B)** 56.1% of respondents meet with 2+ people frequently during the semester (Supplementary S1), and 90.1% of them wore a mask during these gatherings most, if not all, the time. **C)** Most causes of personal noncompliance relate to mask discomfort, specifically poor breathability and thermal regulation.

To understand possible reasons behind negative attitudes towards mask use, a second anonymous survey (n = 536) was sent to Harvard undergraduates. Combined with the results from the first survey, those who noted unfavorable features of masks typically pointed to mask discomfort as the source of their noncompliance (n = 688) (Figure 1C; Supplementary S2). Of note, 68.3% of the reasons for discomfort were issues arising from poor breathability and thermal regulation (i.e., glasses fogging, heat accumulation, trouble breathing, and moisture accumulation). Thus, while compliance is generally high on campus, mask discomfort (particularly issues centered on breathability and thermal regulation) was the primary source of noncompliance.

### Identifying and Validating Key Parameters for Comfort

Because discomfort was found as the primary source of noncompliance, we aimed to identify factors influencing user perception of comfort. Though multiple factors define comfort, the survey highlighted three critical features: air resistance, water vapor permeability, and face temperature change. These parameters influence the breathability and thermal regulation of a mask, which have been considered to be the most significant contributors to mask comfort in the literature (Lee et al, 2020).

#### Air Resistance

Respiratory rate tests of volunteers wearing different types of masks (i.e., surgical/procedural, cotton, and KN95) were conducted to provide physiologic context for the masks’ impact on breathing. First, participants were asked to rate the overall comfort of each mask (Figure 2A). When the comfort ratings were plotted against air resistance values from the literature (Lee et al., 2020; Kim et al., 2015), we found a strong correlation (R^2^ = 0.417) (Figure 2A). With increasing air resistance values, there was a corresponding decrease in perceived comfort as indicated by the increasing comfort scores. Comparison of experimentally determined changes in respiratory rate while wearing various masks relative to the literature air resistance values (Figure 2B) revealed a moderate correlation (R^2^ = 0.256). Increased air resistance was associated with increased average change in respiratory rate.

**Figure 2.**
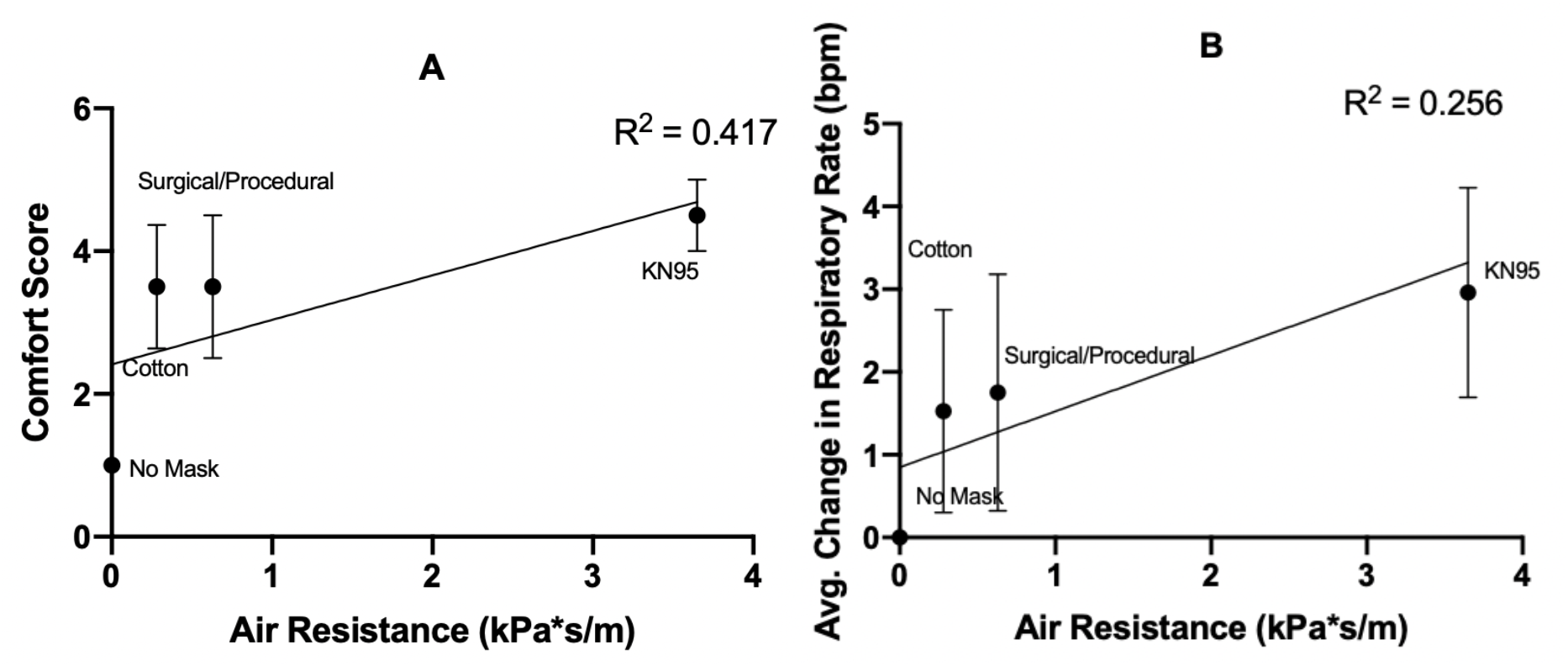
Impact of different masks on perceived comfort and respiration. **A)** Air resistance (kPa*s/m) for all masks plotted against the perceived comfort ratings from 1-10, most to least comfortable (R^2^=0.417 for all points; values represent mean and s.d.). **B)** Average change in respiratory rate (beats per minute) against air resistance (kPa*s/m) of the face masks tested across all participants (n=3 for KN95, n=9 for all others) (R^2^=0.256 for all points; values represent mean and s.d.).

#### Water Vapor Permeability

To address survey reports of moisture accumulation inside masks, the next parameter considered was water vapor permeability. Water vapor permeabilities for three types of filter media (i.e., surgical/procedural, cotton, N95 masks) were collected from literature (Supplementary S3). Cotton masks demonstrated the highest water vapor permeability, while N95 masks demonstrated the lowest. When the participants’ comfort ratings for these masks were plotted against the water vapor permeabilities, there was a moderate correlation (R^2^ = 0.223) (Figure 3). With worsening water vapor permeability (i.e., less negative values), there was also a general decrease in perceived comfort as indicated by an increase in comfort scores.

**Figure 3.**
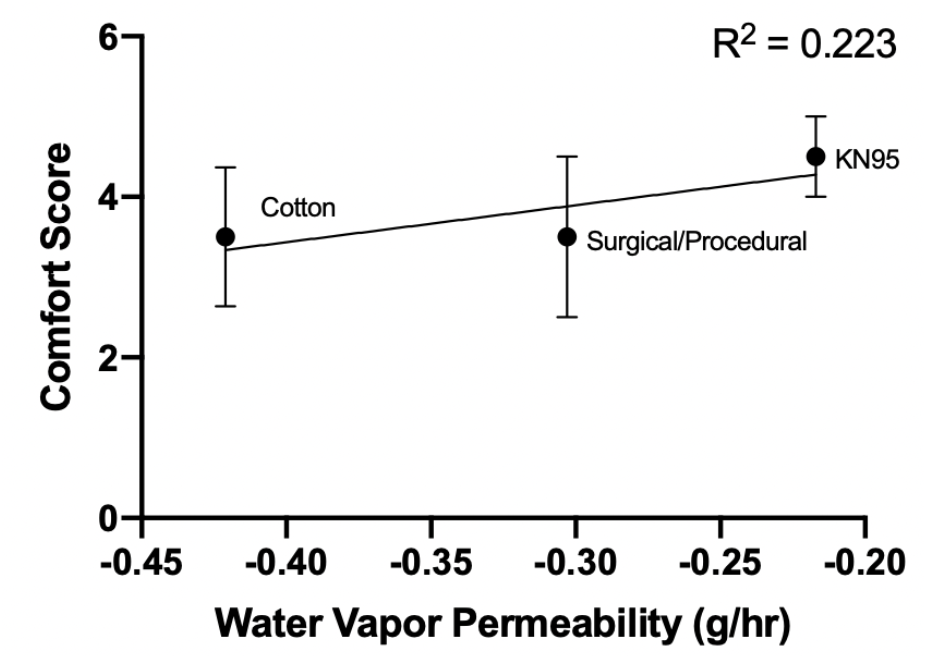
Water vapor permeability (g/hr) for all masks plotted against the perceived comfort ratings (1-10, most to least comfortable) (R^2^ = 0.223 for all points; values represent mean and s.d.).

#### Face Temperature Change

Finally, we analyzed the thermal properties of common masks. The change in facial surface temperature is directly related to a mask’s thermal conductivity. The facial surface temperature of the participants was measured before and after wearing a mask for ten minutes, while either at rest or after exercise (Supplementary S5; Figure 4). During both rest and exercise, surgical/procedural masks led to little warming of the face, similar to not wearing a mask. Cotton masks had a slight insulating effect after exercising as compared to the control group (Figure 4B). In both scenarios examined, KN95 masks significantly increased the facial surface temperature (Figure 4).

**Figure 4.**
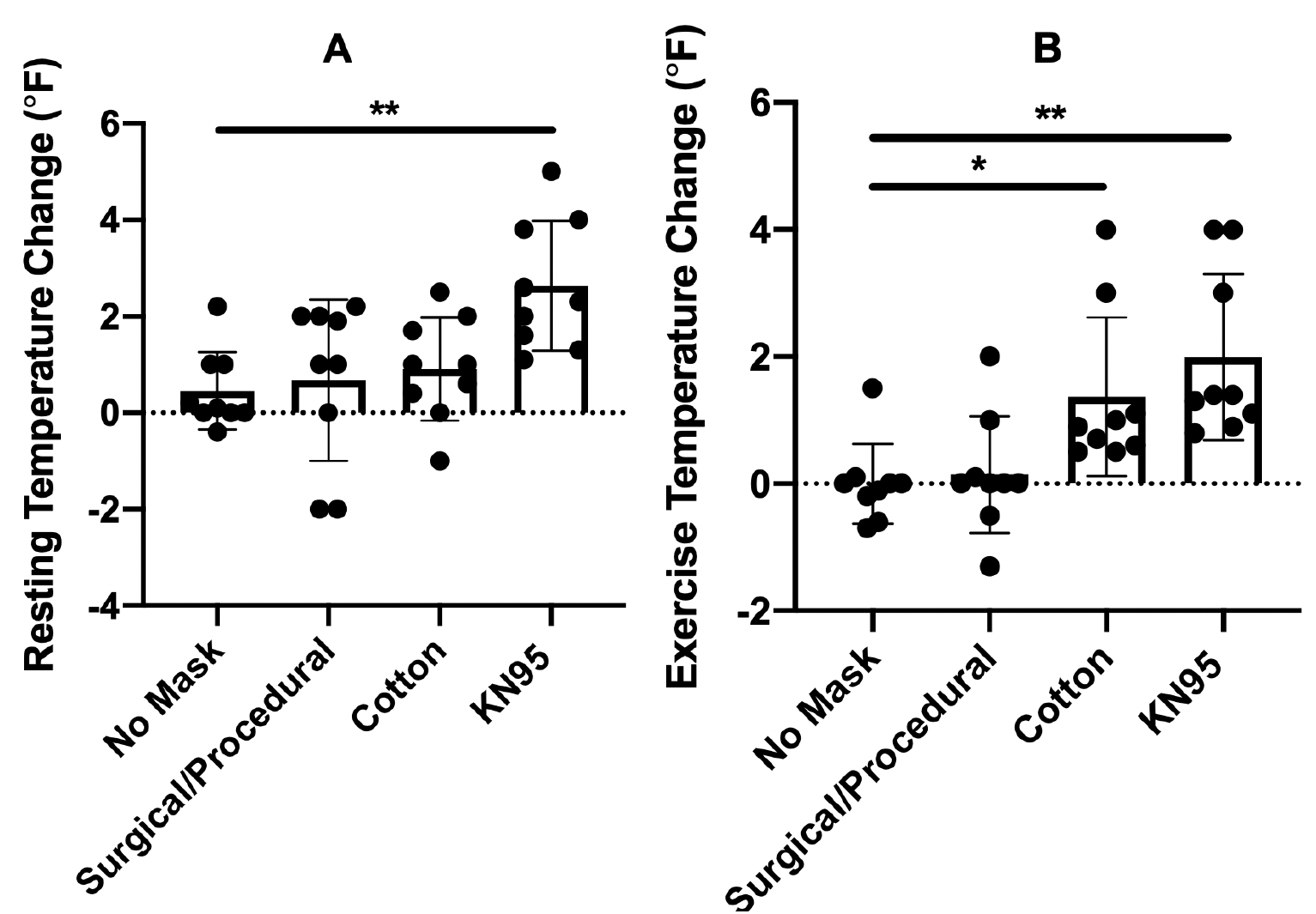
Face temperature change with wearing of different masks. Three trials were performed for each for 3 different participants (n = 9). **A)** Temperature change over 10 minutes at rest (** = p << 0.05). **B)** Temperature change after 10 minutes of exercise (* = p < 0.05, and ** = p << 0.05).

Participants’ comfort rankings (1-10 with 10 being the least comfortable) increased from the initial rating for all masks both while at rest and after exercise (Figure 5A-B). There was a weak correlation between the average comfort ratings for all masks and average temperature changes at both rest and after exercise (R^2^ = 0.066 and 0.193, respectively) (Figure 5C-D). While there was a general deterioration in perceived comfort with greater temperature changes, this relationship appeared to be weak.

**Figure 5.**
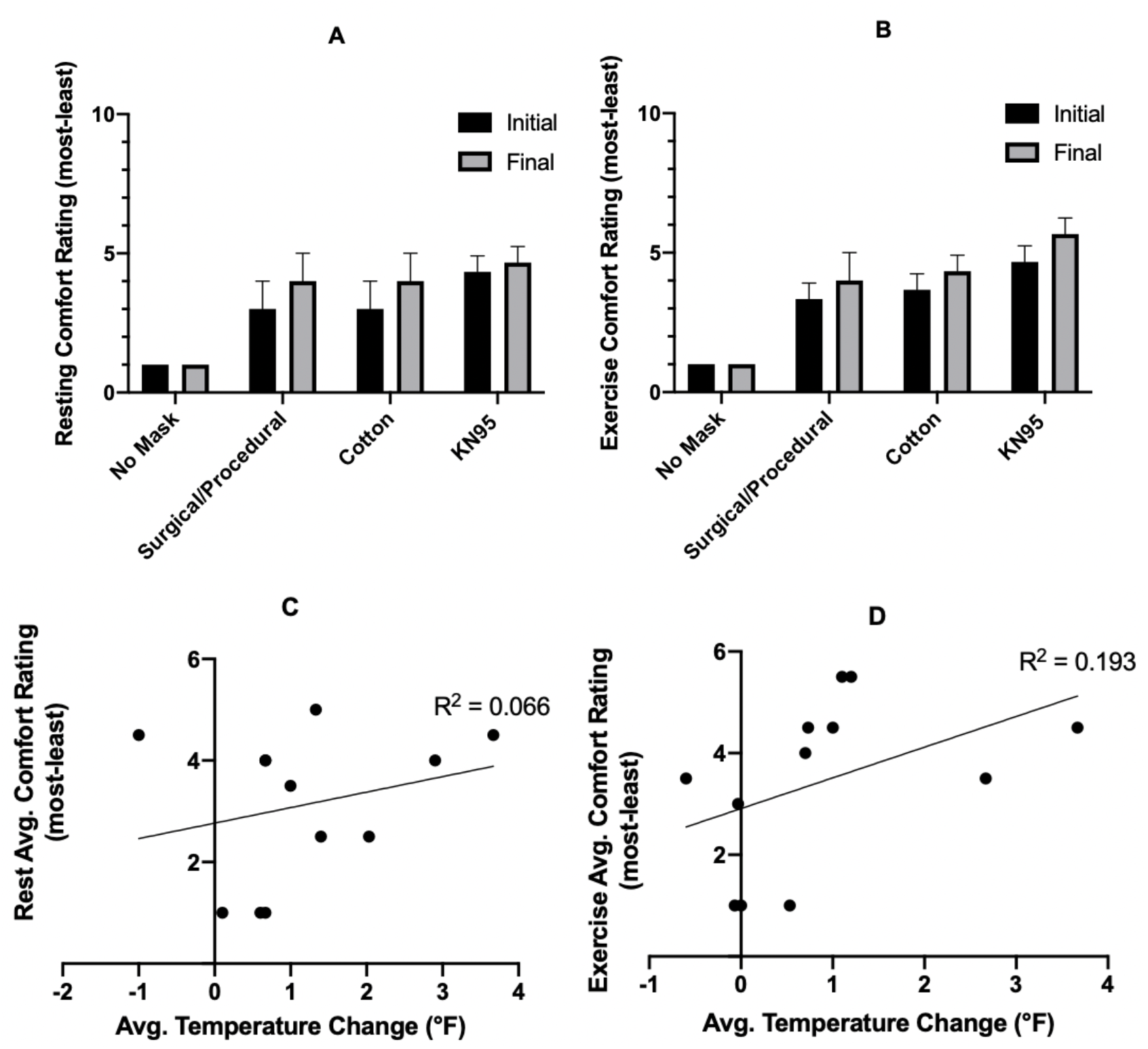
Comfort rating, with 10 being the least comfortable, both initially and after wearing a mask for 10 min. **A-B)** Comfort ratings for each mask tested for the resting (A) and exercise cases (B). Values for all masks showed a statistically significant difference as compared to no mask control. **C-D)** Average temperature change with wearing of all masks plotted against the average perceived comfort ratings during rest (C) and after exercise (D) (R^2^ = 0.066 and 0.193, respectively). Data points represent individual participants and specific masks.

#### Comfort Rankings from Harvard University Campus Surveys

The comfort of various masks was also evaluated through a Harvard University campus survey. When asked to rank the listed face masks (i.e., cotton, surgical/procedural, knitted, polyester, and/or N95/respirators) on a scale of 1-5, with 1 being the most comfortable, survey respondents generally ranked surgical/procedural and cotton masks more favorably (Figure 6; Supplementary S6). Among participants who ranked surgical/procedural masks, 79% scored these masks a 1 or 2 (Figure 6A). Similarly, among participants who ranked cotton masks, 81% scored these masks a 1 or 2 (Figure 6B). In contrast, 42% of respondents ranked N95/respirators at the lower end of the comfort spectrum (Figure 6C). These results match the ranking of these masks according to air resistance, water vapor permeability, and face temperature change (Supplementary S3). This dataset was next utilized to train machine learning algorithms to predict mask comfort.

**Figure 6.**
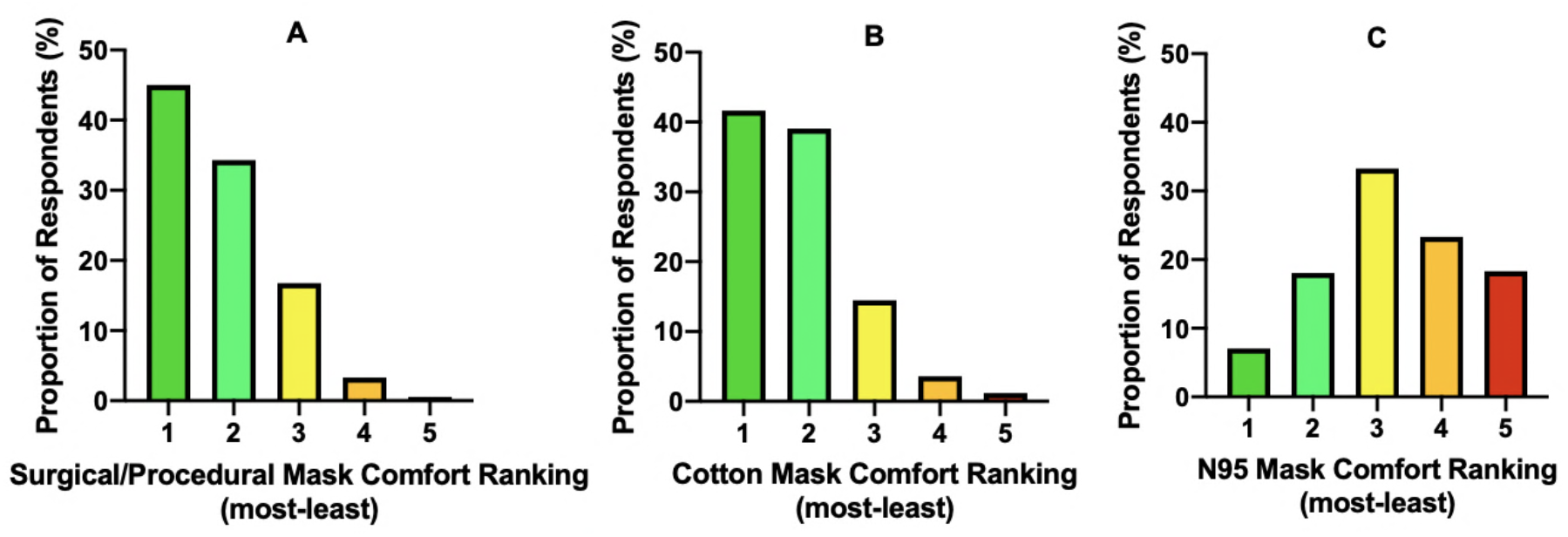
Distribution of comfort rankings for Surgical/Procedural masks **(A)**, Cotton Masks **(B)**, and N95 masks **(C)** by survey participants (1 = most comfortable, 5 = least comfortable). n = 582 for (A); n = 513 for (B); n = 384 for (C).

### Machine Learning (ML) Model Performance and Application

We tested three different ML algorithms to determine the optimal machine learning model to predict mask comfort: a multiple linear regression model (linear regression), random forest, and neural network. The performance of all three models in predicting the comfort score obtained from the survey was fairly similar when given values for the three key parameters (i.e. air resistance, water vapor permeability, and face temperature change) (Table 1).

**Table 1.** Final performance metrics for the three models tested.

|  | Linear Regression | Random Forest | Neural Network |
| --- | --- | --- | --- |
| **Accuracy** | 70.3 % | 70.4 % | 70.4 % |
| **R<sup>2</sup>** | 0.35 | 0.35 | 0.38 |
| **Mean Absolute Error** | 0.84 | 0.84 | 0.84 |
| **Root Mean Squared Error** | 1.04 | 1.04 | 1.02 |

The three models also demonstrate similarity in prediction patterns, as evidenced by visual inspection of graphs of actual and predicted comfort scores, plotted against each of the three parameters, for each model (Figures 7, 8, and 9).

**Figure 7:**
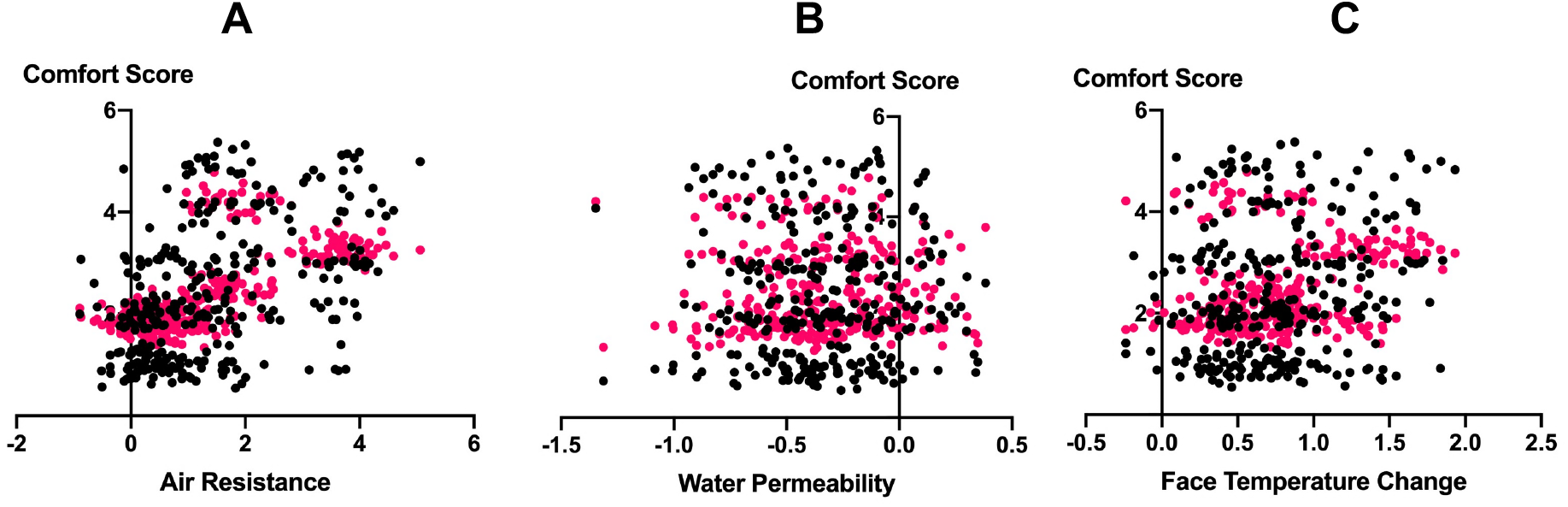
Comfort score, both actual scores (black) and scores predicted by the linear regression machine learning model (pink), plotted against air resistance **(A)**, water permeability **(B)**, and face temperature change **(C)**. These plots are shown with jittered values for ease of viewing. The linear regression model creation and analysis was performed on original un-jittered data.

**Figure 8.**
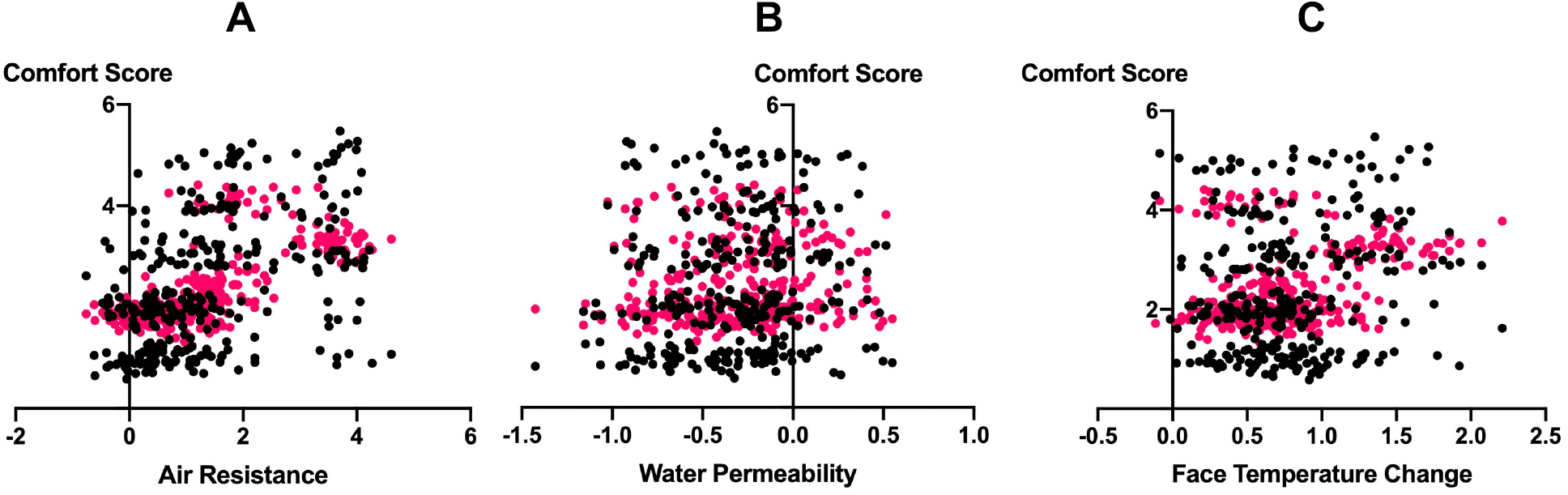
Comfort score, both actual scores (black) and scores predicted by the random forest machine learning model (pink), plotted against air resistance **(A)**, water permeability **(B)**, and face temperature change **(C)**. These plots are shown with jittered values for ease of viewing. The random forest model creation and analysis was performed on original un-jittered data.

**Figure 9.**
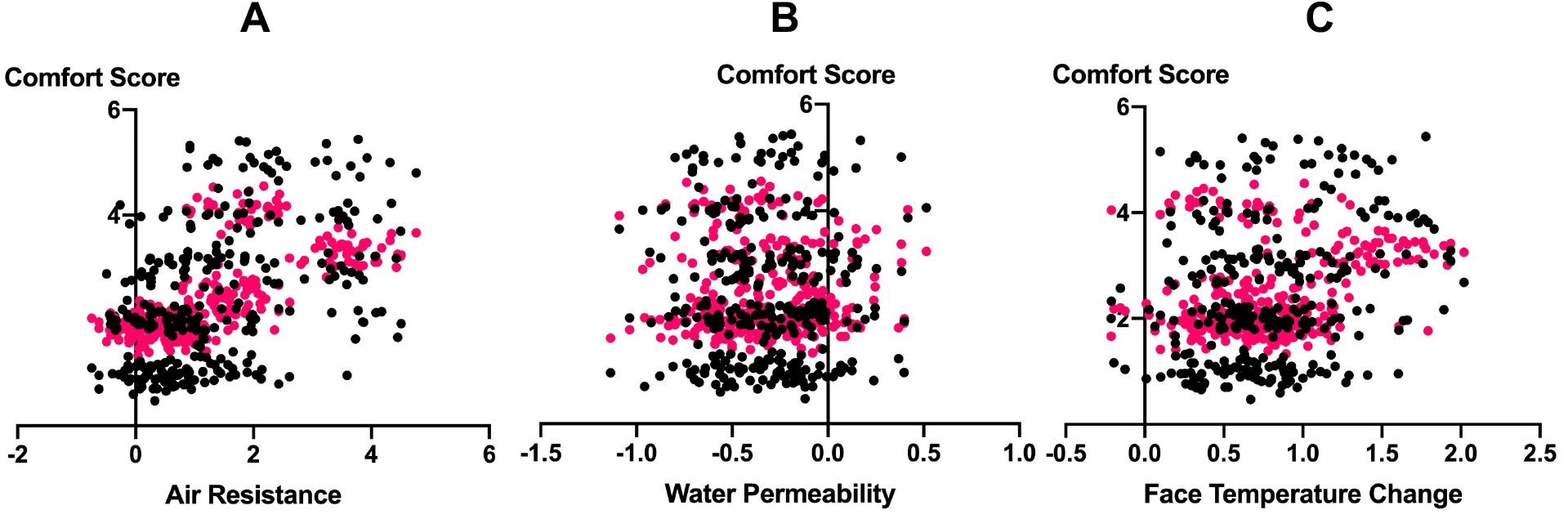
Comfort score, both actual scores (black) and scores predicted by the neural network machine learning model (pink), plotted against air resistance **(A)**, water permeability **(B)**, and face temperature change **(C)**. These plots are shown with jittered values for ease of viewing. The neural network model creation and analysis was performed on original un-jittered data.

All three models predicted comfort scores with approximately the same degree of accuracy (∼70%). Compared to the other two models, the linear regression model offers an advantage in that the functional relationship between the comfort score and the three parameters that is used by the model is readily accessible. This functional relationship provided by the linear regression model follows the functional form below:

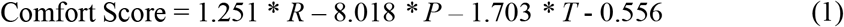

* *Where R is air resistance in units of kPa * s/m, P is water vapor permeability in units of g/hr, and T is face temperature change in units of degrees C*

A 3D visualization of this relationship between the comfort score and the three parameters was created (Figure 10).

**Figure 10:**
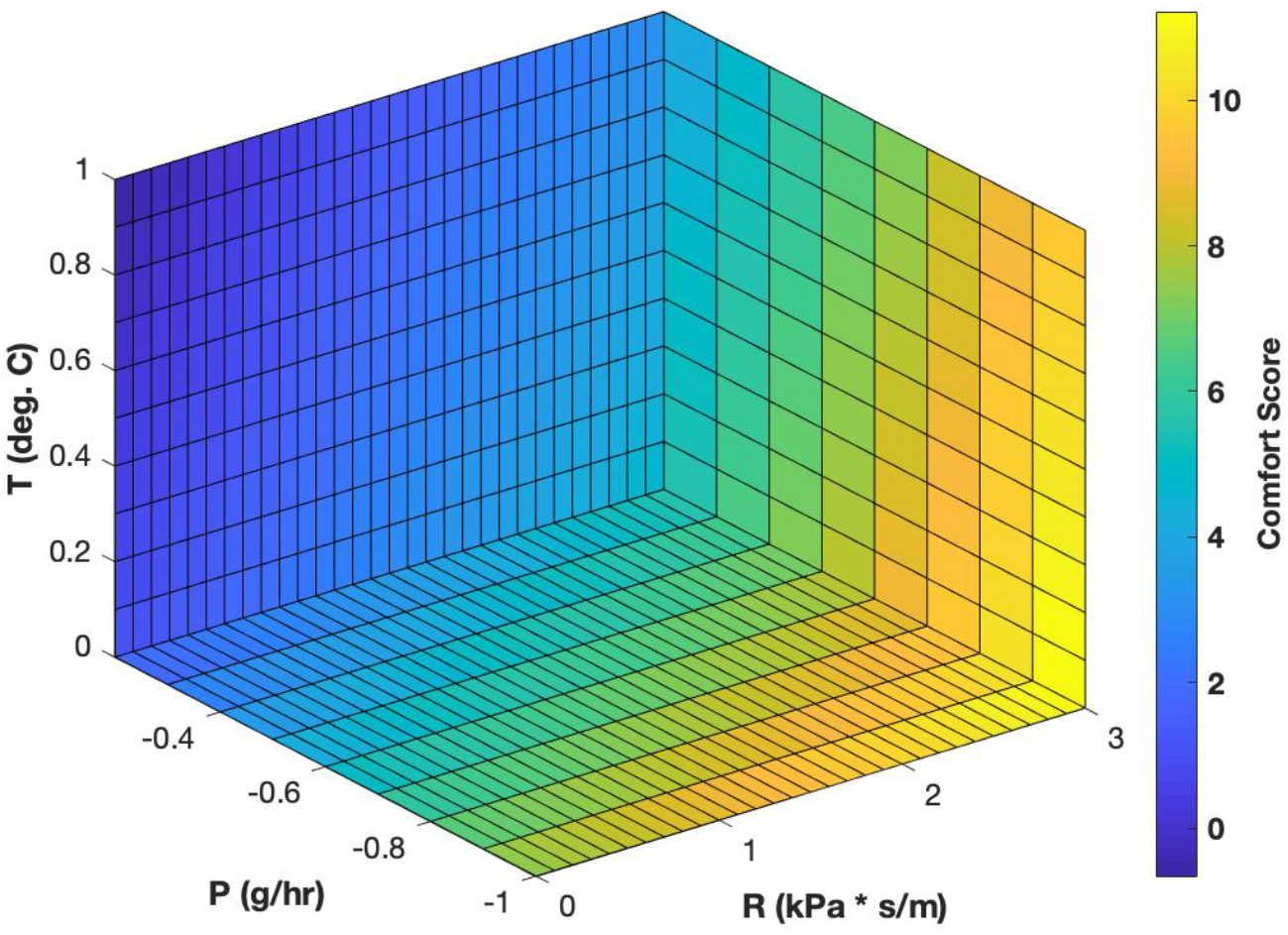
3D visualization of the relationship between comfort score and R (air resistance in kPa * s/m), P (water vapor permeability in g/hr), and T (face temperature change in deg. C).

Using this functional relationship, we analyzed the relative importance of each of the three parameters to the overall perceived comfort of a mask. All three parameters had statistically significant relationships with the comfort score (p < 0.001) (Supplementary S7). Furthermore, the relationship between air resistance and the comfort score was found to have the strongest relationship out of the three parameters (R^2^ = 0.22), while water vapor permeability and face temperature change had weaker relationships (R^2^ = and R^2^ = 0.05, respectively) (Supplementary S7).

#### Application

Finally, we applied our linear regression model to cotton, surgical/procedural, and KN95 masks. As expected, the model predicted a better comfort score (lower value) for the cotton and surgical/procedural masks compared to the KN95 mask (Figure 11).

**Figure 11:**
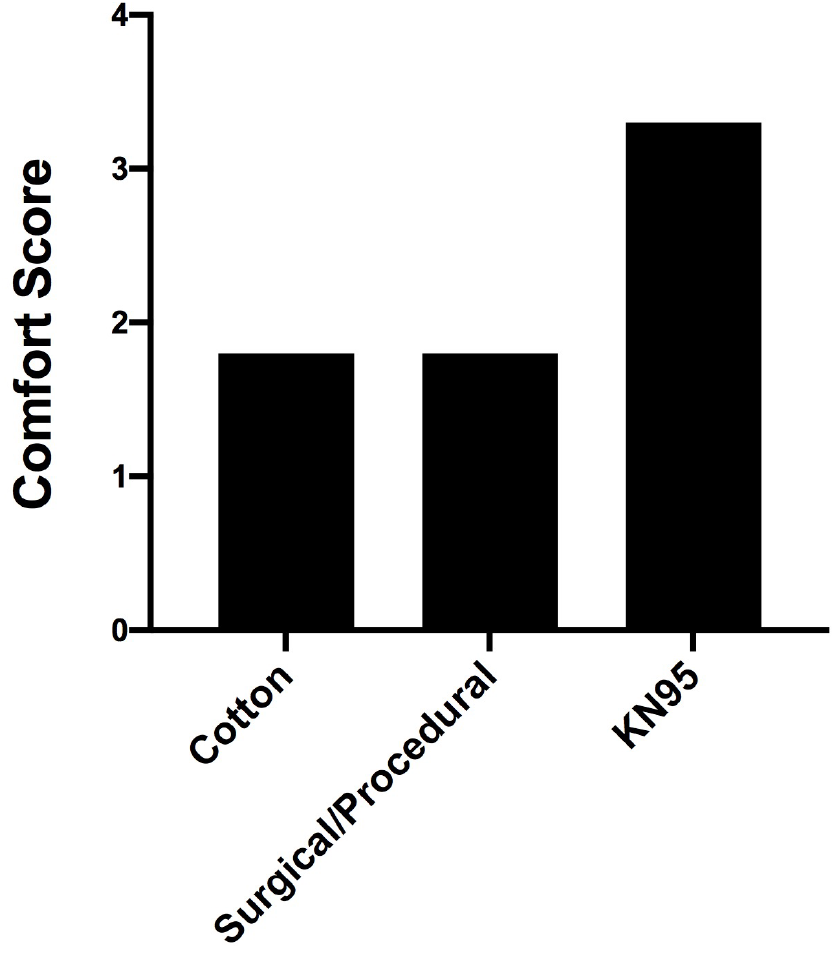
Comfort scores predicted by the linear regression model for control group masks (Cotton - 1.8, Surgical/Procedural - 1.8, KN95 - 3.3). See Supplementary S3 for values used as input parameters in the model.

The comfort scores predicted by this model were close to the true average scores in the survey for these masks (differed by a maximum of ∼0.04) and fall within the 95% confidence interval (Supplementary S8), reinforcing confidence in this model’s utility in quantifying the comfort of a face mask.

## Materials & Methods

### Campus Surveys

#### Compliance

To examine the relationship between comfort and compliance in mask use, the first part of a two-part anonymous survey was conducted. The first part was sent out Fall 2020 to students, staff, and faculty residing on the Harvard campus, and collected compliance data as it related to all the campus mitigation strategies (n=205). The data collected was analyzed by question (Supplementary S10).

#### Ranking Masks for ML

The second anonymous survey collected data regarding campus residents’ perceived comfort of various masks. Only some respondents (n=186) answered the question ranking the comfort levels of masks in the first compliance survey. Respondents to this second part of the survey (n=474) were all undergraduates affiliated with Harvard College who were living both on and off campus at the time the survey launched. Respondents to both surveys (n=660) were students, faculty, and staff at Harvard University. Respondents were not compensated for their participation.

Respondents were asked to rank from MOST to LEAST comfortable the following mask types: cotton, surgical/procedural, N95s (or other respirators), knitted, and polyester. A visual example of each mask type was provided (Supplementary S9). It was not compulsory for respondents to rank all five masks; respondents ranked only the masks with which they had personal experience.

All responses were compiled to quantify relative mask rankings. Masks ranked most comfortable were assigned a value of 1, and rankings were sequentially assigned all the way up to the value of 5 for the least comfortable mask. Finally, for a given mask, the percentage of responses at each rank was calculated over the total number of responses ranking that mask. The ranking calculations were repeated for all five mask types to uncover correlations between perceived comfort and known comfort parameter values across different mask types.

### Parameter Data Collection

#### Comfort Testing Participants Demographics

Eleven subjects conducted mask evaluations to evaluate the efficacy of the comfort model. Eight participants were male, and three participants were female; participants were aged 20 to 31.

#### Masks Used

Subjects evaluated cotton, surgical/procedural, and KN95 masks during testing. For a comparative control, data was also collected from subjects wearing no mask. Participants used their own cotton masks, so brands and sources varied. Procedural masks were 3-ply surgical face masks sourced from Dongguan Weikang Sanitary Products Co., Ltd. in Guangdong, China. KN95 masks were AOK Model #910 manufactured by Shenzhonghai Medical Inc. sourced from Amazon.

#### Water Vapor Permeability

Water vapor permeability parameter data was collected from two publications. The first source, Lee et al. (2020), used ASTM E96 standards to provide data for all face masks except KN95.

Data from Li et al. (2006) was used to approximate the KN95 water vapor permeability from N95 parameters by equating the ratio of the N95 water vapor permeability over the surgical/procedural mask’s water vapor permeability from this study to the ratio of the unknown KN95 water vapor permeability over the surgical/procedural mask’s water vapor permeability from Lee et al. (2020).

#### Air Resistance Values

Data for the air resistance parameter was obtained from existing publications. Lee et al. (2020) determined values in accordance with ASTM D737 guidelines to obtain air resistance (i.e., air permeability) of all five mask types. Supplemental data from Kim et al. (2015) was then used to determine the maximum possible air resistance for N95 masks.

#### Respiratory Rate Test

Vernier Go Direct^®^ respiration belts, alongside Graphical Analysis™ software, were used to measure volunteers’ respiratory rates while under sedentary or active conditions. Volunteers placed the respiratory belt around their upper abdomen, just below the sternum (as per the manufacturer’s instructions), and sat facing away from the monitor to minimize conscious control of breathing. Volunteers then recorded ten seconds each of normal breathing, rapid breathing, and deep breathing, to ensure data was recorded correctly.

Two sets of exercises, normal breathing and accelerated breathing, were collected while wearing each of the following masks: none, procedural, cotton, and KN95 (eight total tests). For normal breathing, volunteers’ breathing rates were recorded for 10 minutes while they were sitting down and reading. For accelerated breathing, volunteers’ breathing rates were recorded for 10 minutes after completing 30 jumping jacks. After the initial 10 minutes, another 50 jumping jacks were performed and breathing was recorded for another 10 minutes. In between the completion of each series of exercises, volunteers were asked to take 20-minute breaks during which eating was not permitted.

#### Face Temperature Test

The face temperature test was performed simultaneously with the respiratory rate test. Prior to testing, all masks were kept at room temperature. During testing, all measurements were collected in a room with constant temperature and humidity. Before the normal breathing respiratory exercise, volunteers were instructed to take three infrared temperature measurements (on the left cheek, below the mouth, and on the right cheek). Immediately after respiratory data was recorded, the volunteers removed their masks and re-collected three temperature measurements for the same previously mentioned facial locations. All measurements collected were within regions covered by any mask. These steps were performed for each mask used for the respiratory rate test.

### Machine Learning Model Development

The goal was to create an algorithm capable of predicting an overall comfort score when given three key parameters: air resistance, water vapor permeability, and face temperature change. Three algorithms were selected based on their levels of complexity: neural network, linear regression, and random forest. Neural networks are complex black box models where the rationale or function for making predictions is not visible to the user. Linear regressions are simple models that result in an end function that is easily accessible, facilitating an in-depth function analysis and extrapolation to other contexts. Random forest models do not provide a functional form describing the method of prediction but provide an intermediate level of complexity.

Published values for air resistance and water vapor permeability were obtained for surgical/procedural, cotton, polyester, knitted, and N95 masks from Lee et al. (2020), Kim et al. (2015), and Li et al. (2006). Published values for face temperature change were obtained for surgical/procedural, cotton, polyester, and knitted masks from Lee et al. (2020). Data obtained for KN95 masks through the face temperature change test described previously was used as a reasonable approximation of the face temperature change for N95 masks, as data for N95 masks is unavailable in Lee et al. (2020). These values served as parameter value inputs in the machine learning models. The comfort rankings of these different masks in the campus survey were used as ground truth comfort scores to be output by the model.

The data was split between training and testing datasets. After collecting all the survey results, there were approximately 660 responses. Since each respondent ranked multiple masks, this amounted to 2046 total data points. After testing different splitting ratios, we found that an 85% training-15% testing split provided a good balance between training accuracy and testing dataset size.

#### Accuracy Metric

A min-max accuracy was chosen as the accuracy metric and is calculated as follows (Prabhakaran, 2016):

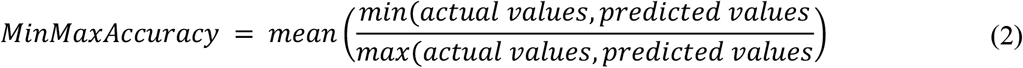

For each data point, the min-max accuracy calculates the ratio of the lower value to the higher value between the actual and predicted values. These ratios are averaged to obtain the final accuracy. In the best-case scenario, if the actual and predicted values are identical, the higher and lower values between the two would be identical, resulting in a ratio of one and a min-max accuracy of 100%. If the actual and predicted values are not identical but are close together, the ratio will be close to one, resulting in a high min-max accuracy, albeit less than 100%.

#### Model Development

The Caret package in R (Kuhn, 2019) was used to create the linear regression and random forest models; the Keras package in R (Falbel et al.) was used to create the neural network model. The neural network had hidden layers with 256 and 128 nodes. The connection between the input layer and the first hidden layer as well as the connection between the first and second hidden layers used the ‘relu’ (rectified linear unit) activation function, while the connection between the second hidden layer and the output layer used a linear activation function. The loss metric used was mean squared error, and the ‘Adam’ optimizer was used with a learning rate of 0.001 and 50 total epochs. The random forest model was created using the root mean squared error as the loss metric. The resampling method was 10-fold cross validation, repeated 3 times, and the final number of variables used for splitting at each node in the final model was 2. The linear regression model was created using the root mean squared error as the loss metric. The resampling method was 10-fold cross validation, repeated 3 times. We also included the possibility of an intercept as a parameter in the model. Each of the models was trained and tested using values for air resistance, water vapor permeability, and face temperature change as independent variables and comfort ratings from the survey as the dependent variable.

### Statistical Analyses

Statistical analysis of comfort parameter data was performed using GraphPad Prism v8.4.3. Copyright 2020 GraphPad Prism is a registered trademark of GraphPad Software, LLC. All values of n can be found in the Results section and refer to the number of human subjects. Comparison of face temperature changes between tested masks were performed using one-way ANOVA. Normality was checked using the D’Agostino & Pearson test. A p value of <0.05 was considered significant.

Machine learning model development and performance analysis was performed using RStudio v.1.2.5033 and associated packages. Copyright 2009-2019 RStudio is a registered trademark of RStudio, Inc. Significance of relationships between each parameter and comfort score was determined in GraphPad Prism using the F-test. A p value of <0.05 was considered significant.

## Discussion

Survey results largely point to mask discomfort as the underlying reason behind personal noncompliance with mask donning policies. Mask breathability and thermal regulation are considered the most significant attributes impacting comfort. Current models of comfort frequently rely on qualitative ratings or quantitative assessments using a single comfort parameter. In contrast, our machine learning model predicts a quantitative comfort score that accounts for three of the most influential parameters involved in perceived mask comfort

Unlike past surveys gauging respirator comfort among healthcare workers (Baig et al., 2010; Krah et al., 2016), this study reports a large survey comparing different face coverings in a non-healthcare setting. Among university affiliates, user compliance was largely dominated by mask comfort. Thus, it is important for masks to not only be effective at filtering particles but also comfortable in order to mitigate disease transmission.

The comfort parameters we used to capture mask breathability and thermal regulation were air resistance, water vapor permeability, and face temperature change. These core parameters were examined in past work (Lee et al., 2020). Goodness-of-fit (i.e., the ability of a mask to fit the facial contours) was also identified as an important factor of comfort (Lee et al., 2020). However, this parameter was qualitatively evaluated in Lee et al. and was not considered in our study to avoid cross-interactions with air resistance as better fit may lead to greater air resistance. Other studies seeking to evaluate mask comfort rely on qualitative surveying of comfort rather than on a detailed, quantitative study of various comfort properties (Baig et al., 2010; Krah et al., 2016; Wentworth et al., 2021). The ASTM’s focus on ensuring mask breathability is consistent with our identification of air resistance as the most significant comfort parameter. Comfortable barrier face coverings abiding by ASTM standards should exhibit air resistances lower than 1.56 kPa*s/m (ASTM, 2021).

Other comfort parameters, such as mask rigidity and mass distribution, were not analyzed but may be useful to consider. ASTM standards for barrier face coverings specify that materials that touch the skin must be non-irritating, nontoxic, and free of sharp edges, points, or burrs (ASTM, 2021). In such cases, frame rigidity could correlate with comfort since highly rigid masks may dig into a wearer’s skin, a frequently reported cause of discomfort in our campus survey. Mass distribution may also impact comfort as a mask would feel heavier if the center of gravity is farther from the face.

Our linear regression model has approximately 70% accuracy and provides a functional form for the relationship between the parameters and the predicted comfort score. The model’s predictions falling within the 95% confidence interval of the true campus survey mean comfort score for all five masks in the survey and the narrow margin between average survey rankings and predicted comfort scores provide strong evidence of the model’s ability to accurately predict comfort scores (Supplementary S8). The functional form of the relationship between the parameters and the comfort score also reveals that air resistance has a much greater numerical contribution to the comfort score than face temperature change and water vapor permeability (Supplementary S7). Nevertheless, all three parameters significantly contribute to the overall comfort score and are therefore essential components of the function (Supplementary S7). These results are supported by trends observed during participant testing, in which participant perception of mask comfort correlated more strongly with air resistance than water vapor permeability or face temperature change (Figures 2A, 3, 5).

A deep learning model has been previously developed to detect whether an individual is wearing a face mask (Loey et al., 20201). Additionally, Support Vector Machine and Ensemble Algorithm models have been tested to predict comfort in other contexts, such as personal thermal comfort modeling (Katić et al., 2020). Similar to the machine learning model of mask comfort we have presented, these models demonstrate intermediate accuracy, as is expected given the subjective nature of the predictions. These models also rely on more complex algorithms than our linear regression model, as parameters involved in object detection and broader personal thermal comfort assessment are likely nonlinear. Although these approaches exist to implement machine learning as a prediction technique for mask detection and broad personal thermal comfort assessment, to the best of our knowledge, machine learning models have not yet been created for predicting personal comfort while wearing different face masks.

## Conclusions

We describe a quantitative model to gauge face mask comfort using three parameters: air resistance, water vapor permeability, and face temperature change. This model allows users to assess the relative comfort of a mask and choose one that best suits their comfort needs. Its use is expected to minimize discomfort-induced noncompliance. As mask-wearing is also essential in contexts outside of SARS-CoV-2 transmission mitigation, including several industrial settings, this model is likely to provide a useful tool in many scenarios.

## Data Availability

Data is available upon request.

## Acknowledgements

We would like to acknowledge Nicholas Jeffreys, the Harvard Face Mask Committee, and the Harvard Active Learning Labs for providing excellent guidance throughout our investigation process. We would especially like to acknowledge Harvard Face Mask Committee members Stephen Blacklow, John Doyle, Willy Shih, Mary Corrigan, Sarah Fortune, and Sara Malconian for their invaluable support and advice. We would also like to recognize Evan Hunsicker and Katia Osei for their initial data analysis, and Paola Carrillo Gonzalez, Jordan Daigle, Taisa Kulyk, Kazi Tasnim, Meghan Turner and James Young for their contributions during the Harvard SEAS engineering design course. Last but not least, we would like to thank Juan C., Mason D., Benjamin F., Rosie P., Daniel R., Miguel S., and Yann T. for volunteering as additional study participants.

