## Supplemental Information for "Quantifying Face Mask Comfort"

### Supplementary Information

**S 1. Compliance of Mask Use in Gatherings**

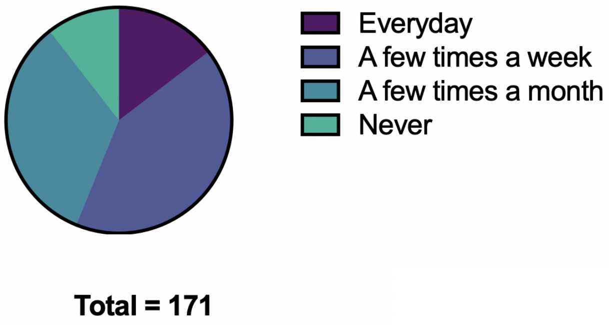


**Figure S1.** Compliance of mask use of survey participants when attending gatherings of 2+ people, discounting roommates (n=171).

**S 2. Characteristics of Mask Discomfort (Other)**

**Table S2.** Characteristics of current masks survey participants (n = 688) listed that they disliked.

| What characteristics of current masks do you dislike? - Other |
|---|
| Sometimes my face will get irritated |
| Sometimes light-headed from walking with mask on |
| The earloop strings often break. Have to carry two just in case. |
| moisture from my breath accumulates, which does not cause the mask to stick to my face but is still uncomfortable |
| the masks are too big/need a lot of readjustment |
| It's itchy on my face. Irritates my face |
| Acne caused by mask |
| Causes mask acne |
| Makes my face breakout |
| I run with my mask on, humidity makes it challenging to breath |
| Some surgical masks are too large and dont conform to my face |
| I get acne where the mask touches my skin |
| They are usually way too big on my face. |
| Maskne |
| Curls my beard |
| size can be issue. loops pull on ears and give me headache |
| mask slides down my nose |

| |
|---|
| I don't trust the filtration (or there's no filter at all) |
| Too loose/has gaps |
| Mask Acne |
| Earloops that aren’t adjustable :( |
| exercising |
| Getting acne from masks |
| Hard to exercise in masks |
| The nose size doesn't fit, causing the mask to dig into my nose |
| The extent to which masks mold to the jawline is very awkward. Instead of following what is a prominent bone structure of most faces, they approximate it with round or square shapes. |
| Cotton mask feels heavy on my face. I'm happy with/used to my surgical mask though! |
| The current masks make certain exercises difficult like running. |
| Masks are often too big on me and fall off my ears |
| Masks don't fit quite right |
| ACNE |
| The ear loops tend to fall off |
| Trouble breathing only while running |
| Slips down off my nose |
| Hard to talk--mask gets in mouth when it doesnt have enough structure |
| if i'm not careful, i break out on my face where the mask is |
| Some do not confirm to the contours of my face, leaving gaps. |
| Some masks don't cover my entire (short) beard, and I prefer masks that do. |
| the little threads on the surgical masks make me sneez |
| mask isn't a snug fit on my face so it slides down |
| ear loops on fabric masks loosen too quickly (either the elastic stretches out or the knot comes undone) |
| Hard to do work with mask on, difficulty breathing, odors |
| uncomfortable around nose |
| rubber loops on N95s pull on hair, and the ear loops on masks need to be the correct length so they don't yank on your ears, an adjustable length on the ear loops would be a good feature |
| my nose keeps falling out |
| Ones that don’t tie stretch out, leading to falling masks and a constant need for readjustment. |
| Hard to speak |
| acne |

| |
|---|
| I wanna cultivate my neckbeard but the mask makes me look like an ogre |
| I get headaches when I wear masks |
| See previous answer. My issue is both all of the above and much more with respect to principles. |
| makes my nose itchy |
| Scent |
| I don't feel protected enough by certain masks, some leave too much space for air to get in |

**S 3. Key Parameters**

**Table S3.** Key parameter values for different face masks.

| **Material** | **Air Resistance (kPa*s/m)** | **Water Vapor Permeability (g/hr)** | **Face Temperature Change (°C)** |
|---|---|---|---|
| Surgical/Procedural Mask* | 0.63 | -0.303 | 0.5 |
| Cotton Mask* | 0.28 | -0.421 | 0.8 |
| N95 Mask** | 3.65 | -0.217 | 1.4611 |
| KN95 Mask*** | 3.65 | -0.217 | 1.4611 |

** All input parameter values were derived from Lee et al., 2020.*
*** Air resistance and water vapor permeability values were derived from values from Kim et al., 2015 and Li et al., 2006 respectively; face temperature change value was estimated from KN95 mask face temperature tests.*
**** Air resistance and water vapor permeability values were estimated from N95 values; face temperature change value was obtained from face temperature change tests previously described.*

**S 4. Air Resistance**

**Table S4:** Changes in average Respiratory Rate for each participant in breaths per minute observed for various masks

| | | **Change in Average Respiratory Rate for Participant (breaths / min)** | | | | | | | | | |
|---|---|---|---|---|---|---|---|---|---|---|---|
| **Mask** | **Air Resistance (kPas/m)** | **1** | **2** | **3** | **4** | **5** | **6** | **7** | **8** | **9** | **Average** |
| **No Mask** | 0 | 0 | 0 | 0 | 0 | 0 | 0 | 0 | 0 | 0 | 0 |
| **Cotton Mask** | 0.28 | 0.0341 | 3.7522 | 0.4133 | 2.751 | 0.3558 | 1.9122 | 1.3761 | 1.0476 | 2.1093 | 1.5280 |
| **Surgical/Procedural Mask** | 0.63 | 3.0582 | 4.9622 | 1.3923 | 1.9807 | 0.6925 | 0.9903 | 1.2934 | 0.8578 | 0.5323 | 1.7511 |
| **KN95** | 3.65 | 3.0919 | 4.1567 | 1.6349 | NaN | NaN | NaN | NaN | NaN | NaN | 2.9611 |

**S 5. Face temperature change**

**Table S5.** Face temperature increase and comfort level for tested masks.

| **Mask** | **Average Face Temperature Change (°C)** | **Average Comfort Level (1-10, 10 most comfortable)** |
|---|---|---|

| | 10 Minutes Rest | 30 Jumping Jacks | 10 Minutes Rest | 30 Jumping Jacks |
|---|---|---|---|---|
| No mask | 0.3 | 0.1 | 10 | 10 |
| Cotton | 0.5 | 0.8 | 7.5 | 7 |
| Surgical/Procedural | 0.4 | 0.1 | 7.5 | 7.3 |
| KN95 | 1.4 | 1.1 | 6.5 | 5.7 |

**S 6. Distribution of comfort rankings for listed masks by survey participants**

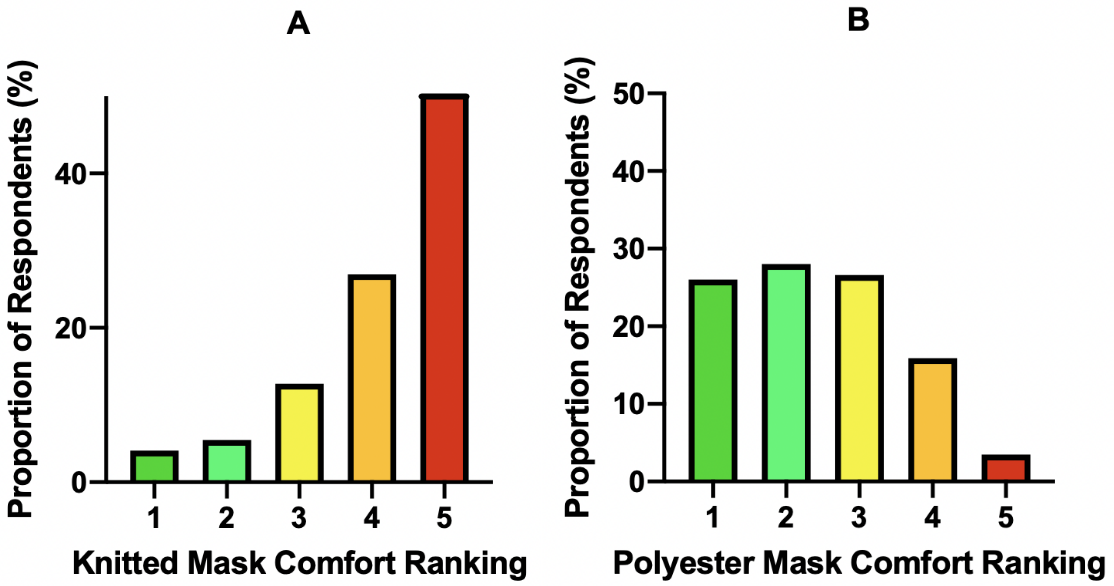


**Figure S6.** Distribution of comfort rankings for listed masks by survey participants (1 = most comfortable, 5 = least comfortable). **A)** 77.58% of participants who ranked knitted masks (n = 219) scored them 4 or 5. **B)** 54% of participants who ranked polyester masks (n = 346) scored them 1 or 2.

**S 7. Correlation between each key parameter to comfort score**

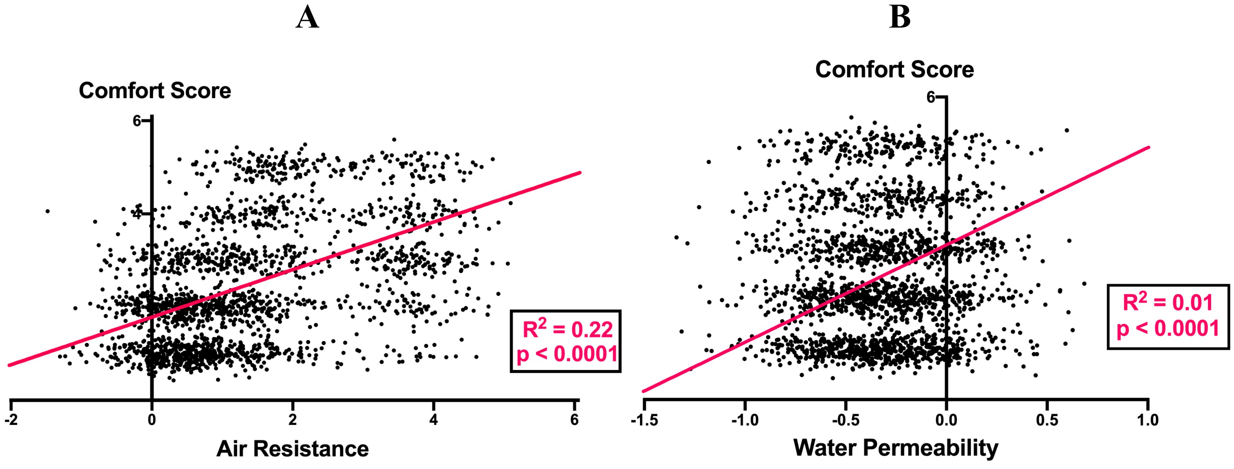

**C**

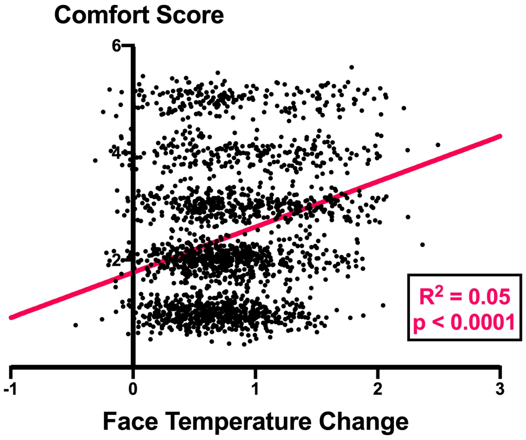


**Figure S7.** Correlation between Comfort Score and **(A)** Air Resistance **(B)** Water Vapor Permeability and **(C)** Face Temperature Change. P values for each parameter were calculated through the F-test.

**S 8. Confidence interval analysis of comfort scores**

| | Cotton Mask | Surgical/ Procedural Mask | Polyester Mask | N95 Mask | Knitted Mask |
|---|---|---|---|---|---|
| **Average Comfort Score in Campus Survey** | 1.80 | 1.84 | 2.44 | 3.28 | 4.15 |
| **95% Confidence Interval** | (1.73, 1.88) | (1.76, 1.91) | (2.31, 2.56) | (3.16, 3.40) | (4.00, 4.30) |
| **Comfort Score Predicted by Linear Regression Model** | 1.81 | 1.81 | 2.48 | 3.26 | 4.16 |

**S 9. Reference figure for survey respondents to rank the comfort of different masks**

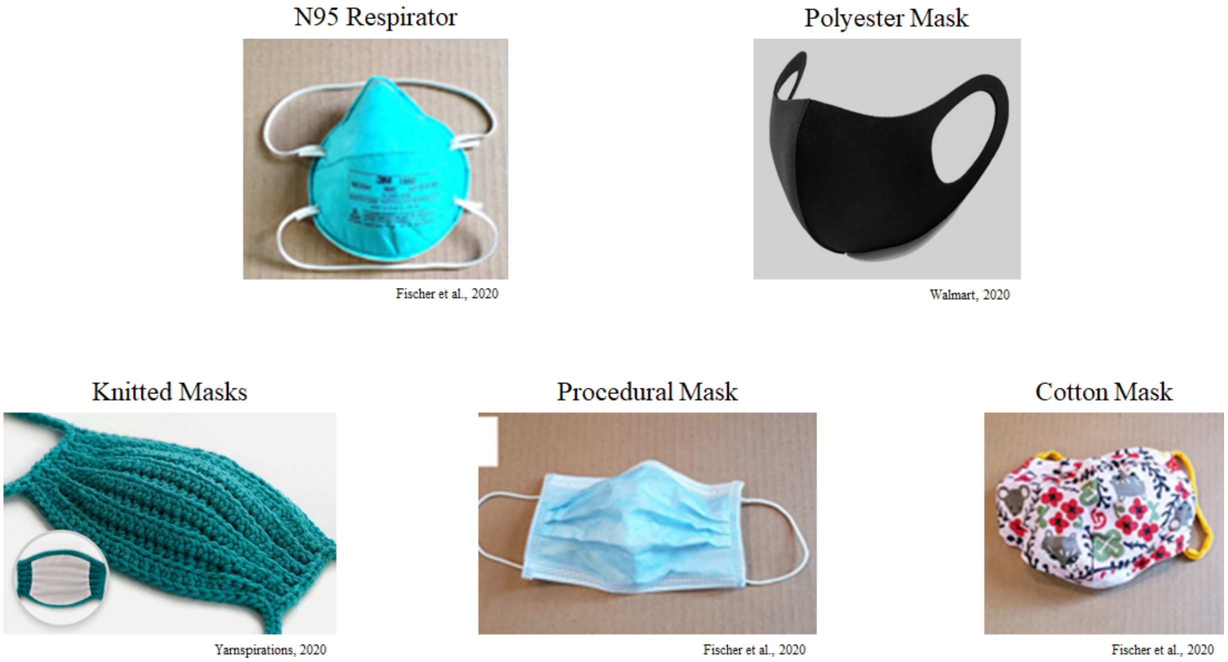


**Figure S9.** Reference figure for survey respondents to rank the comfort of different masks. Example images of cotton masks, surgical/procedural masks, N95s (or other respirators), knitted masks, and polyester masks were provided.

**S 10. Survey Questions**

1. Please rank the below masks and face coverings you've worn from MOST comfortable to LEAST comfortable. (Drag and Drop)

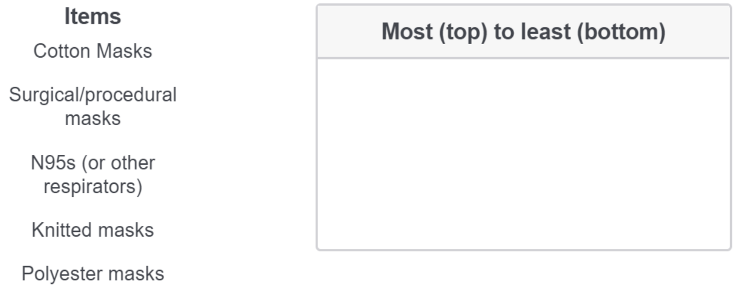
Items
Cotton Masks
Surgical/procedural masks
N95s (or other respirators)
Knitted masks
Polyester masks
Most (top) to least (bottom)

2. What characteristics of current masks do you dislike?
    a. Earloops irritate my ears
    b. I have trouble breathing with my mask on
    c. Masks cause my glasses to fog up
    d. Moisture from my breath accumulates, causing the mask to stick to my face
    e. Heat accumulates inside my mask
    f. Mask frame material digs into my face
    g. None - the current masks are perfect
    h. Other [Open-Ended Text Response]
3. How often do you meet with a group of 2+ people (discounting roommates)?

    a. Everyday
    b. A few times a week
    c. A few times a month
    d. Never
4. For these gatherings of 2+ people:
    a. Do you wear a mask during these gatherings?
        i. Always
        ii. Most of the time
        iii. Sometimes
        iv. Rarely
        v. Never
    b. Do you meet outdoors?
        i. Always
        ii. Most of the time
        iii. Sometimes
        iv. Rarely
        v. Never
    c. If indoors, how often do you have your HEPA filter on?
        i. Always
        ii. Most of the time
        iii. Sometimes
        iv. Rarely
        v. Never
5. On a scale of 1-10, to what extent do you think people on campus follow social distancing rules?
    a. 1: No one follows the rules
    b. 10: Everyone always follows the rules
6. On a scale of 1-10, how likely do you think consequences will be enforced?
    a. 1: No one follows the rules
    b. 10: Everyone always follows the rules
7. How often are your nose or mouth uncovered when wearing a mask?
    a. All the time
    b. Most of the time
    c. Sometimes
    d. Rarely
    e. Never
8. How often do you wear a mask that is NOT provided by the University (i.e. NOT the procedural masks)?
    a. Always
    b. Most of the time
    c. Sometimes
    d. Rarely
    e. Never
9. Do you ALWAYS keep the HEPA filter in your room running?
    a. Yes

    b. No
        i. If no, why not? [textbox]

10. Do you wear a mask while walking/commuting on campus and around the Square?
    a. Always
    b. Most of the time
    c. Sometimes
    d. Rarely
    e. Never
11. If you don’t wear a mask, why not? [textbox]
12. How many non-dining workers do you encounter while picking up your meal?
    a. 0 people
    b. 1-2 people
    c. >3 people
13. Do you wear a mask outside of your dorm room while still in your house?
    a. Always
    b. Most of the time
    c. Sometimes
    d. Rarely
    e. Never
14. How frequently do you dine at non-Harvard cafes/restaurants?
    a. Everyday
    b. A few times a week
    c. A few times a month
    d. A few times a semester
    e. Never
15. How frequently do you eat a meal with one or more people? (discounting roommates)
    a. Everyday
    b. A few times a week
    c. A few times a month
    d. A few times a semester
    e. Never
